# Almost significant: trends and P values in the use of phrases describing marginally significant results in 567,758 randomized controlled trials published between 1990 and 2020

**DOI:** 10.1101/2021.03.01.21252701

**Authors:** Willem M Otte, Christiaan H Vinkers, Philippe Habets, David G P van IJzendoorn, Joeri K Tijdink

## Abstract

**Objective:** To quantitatively map how non-significant outcomes are reported in randomised controlled trials (RCTs) over the last thirty years.

**Design:** Quantitative analysis of English full-texts containing 567,758 RCTs recorded in PubMed (81.5% of all published RCTs).

**Methods:** We determined the exact presence of 505 pre-defined phrases denoting results that do not reach formal statistical significance (P<0.05) in 567,758 RCT full texts between 1990 and 2020 and manually extracted associated P values. Phrase data was modeled with Bayesian linear regression. Evidence for temporal change was obtained through Bayes-factor analysis. In a randomly sampled subset, the associated P values were manually extracted.

**Results:** We identified 61,741 phrases indicating close to significant results in 49,134 (8.65%; 95% confidence interval (CI): 8.58–8.73) RCTs. The overall prevalence of these phrases remained stable over time, with the most prevalent phrases being ‘marginally significant’ (in 7,735 RCTs), ‘all but significant’ (7,015), ‘a nonsignificant trend’ (3,442), ‘failed to reach statistical significance’ (2,578) and ‘a strong trend’ (1,700). The strongest evidence for a temporal prevalence increase was found for ‘a numerical trend’, ‘a positive trend’, ‘an increasing trend’ and ‘nominally significant’. The phrases ‘all but significant’, ‘approaches statistical significance’, ‘did not quite reach statistical significance’, ‘difference was apparent’, ‘failed to reach statistical significance’ and ‘not quite significant’ decreased over time. In the random sampled subset, the 11,926 identified P values ranged between 0.05 and 0.15 (68.1%; CI: 67.3–69.0; median 0.06).

**Conclusions:** Our results demonstrate that phrases describing marginally significant results are regularly used in RCTs to report P values close to but above the dominant 0.05 cut-off. The phrase prevalence remained stable over time, despite all efforts to change the focus from P < 0.05 to reporting effect sizes and corresponding confidence intervals. To improve transparency and enhance responsible interpretation of RCT results, researchers, clinicians, reviewers, and editors need to abandon the focus on formal statistical significance thresholds and stimulate reporting of exact P values with corresponding effect sizes and confidence intervals.

**Significance statement:** The power of language to modify the reader’s perception of how to interpret biomedical results cannot be underestimated. Misreporting and misinterpretation are urgent problems in RCT output. This may be at least partially related to the statistical paradigm of the 0.05 significance threshold. Sometimes, creativity and inventive strategies of clinical researchers may be used – describing their clinical results to be ‘almost significant’ – to get their data published. This phrasing may convince readers about the value of their work. Since 2005 there is an increasing concern that most current published research findings are false and it has been generally advised to switch from null hypothesis significance testing to using effect sizes, estimation, and cumulation of evidence. If this ‘new statistics’ approach has worked out well should be reflected in the phases describing non-significance results of RCTs. In particular in changing patterns describing P values just above 0.05 value.

More than five hundred phrases potentially suited to report or discuss non-significant results were searched in over half a million published RCTs. A stable overall prevalence of these phrases (10.87%, CI: 10.79–10.96; N: 61,741), with associated P values close to 0.05, was found in the last three decades, with strong increases or decreases in individual phrases describing these near-significant results. The pressure to pass scientific peer-review barrier may function as an incentive to use effective phrases to mask non-significant results in RCTs. However, this keeps the researcher’s pre-occupied with hypothesis testing rather than presenting outcome estimations with uncertainty. The effect of language on getting RCT results published should ideally be minimal to steer evidence-based medicine away from overselling of research results, unsubstantiated claims about the efficacy of certain RCTs and to prevent an over-reliance on P value cutoffs. Our exhaustive search suggests that presenting RCT findings remains a struggle when P values approach the carved-in-stone threshold of 0.05.

## Introduction

Individual clinical researchers are subject to regulations, traditions and procedures such as the mythical heritage or paradigm of the peculiar and well recognized 0.05 significance threshold. Individuals submitting RCT publications are dancing the ‘significance dance’ to reach outcomes below the five percent alpha level. This leads to a Catch-22 situation, in particular when calculated P values are just above 0.05. To convince reviewers about the usefulness of data that nonetheless did not reach statistical significance, based on this artificially fixed threshold, is a challenge. Interestingly, the vast majority (96%) of biomedical articles report P values of 0.05 or less (1, 2). Unseen, but behind this peculiar distribution of published P values, are all those P values that did not make it below 0.05. In psychology, the occurrence of reporting P values between 0.05 and 0.1 – about 40% – is relatively high (3). Less is known about these numbers in clinical research. In a small sample of 722 articles in oncology research, 63 articles (8.7%) used trend statements to describe statistically non-significant results (4).

Strong preferences for P values below 0.05 may lead to creative linguistic solutions. Reporting non-significant results as important or noteworthy findings may effectively invite scholars to overstate their findings and present uncertain, low evidence results, as important ‘breakthrough’ research with clear clinical impact. This struggle has an evolutionary element. Some language phrases will be more successful in convincing editors and reviewers than others. A well-known approach is to present non-significant results as pseudo-significance. Given the relative conservative RCT research environment, we expected both creative linguistics regarding significance phrases in published RCTs as well as substantial dynamics over time for the most favourite phrases.

Insight in this practice is important as the success of an RCT is partly determined by the way the results are presented in a manuscript (5). Effective interventions and procedures with clear and significant outcomes that promise to improve patient care will most likely guide the decision on acceptance. However, in papers without clear clinical breakthroughs, the language used to highlight potential beneficial treatments may nonetheless convince reviewers and readers of the importance of the results (6, 7). Also, for RCTs, the cornerstone of evidence-based medicine, two independent meta-analytic studies have detected that positive reporting and interpretation of primary outcomes in RCTs were frequently based on non-significant results (8, 9). Furthermore, Chiu and colleagues report semantic ‘overselling’ of statistically non-significant results and inappropriately use of causal statements are used in approximately half of the inspected 374 RCTs (10). Persuasive phrasing like ‘marginally significant’ and ‘a trend towards significance’, may disguise non-significant results. Given that there is essentially no clinically relevant distinction between a type I error of 4, 5 or 6%, it is interesting to understand how the formulations regarding P values just above 0.05 change over time.

In this study, we therefore quantitatively analysed 567,758 RCT full-texts, registered in the last three decades in the PubMed database. We determined the use of most common phrases describing non-significant results, we characterized the trends over time, and, in a subset, their associated P values. We expected to find similar percentages of phrases associated with non-significant results in RCTs as reported in other (mostly non-clinical) studies (8-10). We also hypothesized to detect changes in phrase prevalences over time, assuming continuous evolution of phrasing in reporting of non-significant RCT results. Finally, we anticipated that the phrase-associated P values would predominantly be associated with a P value in the range of 0.05–0.15.

## Methods

### Selection of RCTs

We identified all RCTs in the PubMed database and excluded animal studies and studies which were not actual RCT reports [Sep 20, 2020]. Subsequently, we collected the Portable Document Format (PDF) for all available RCTs across publishers in journals covered by the library subscription of our institution and we converted the PDFs to structured plain text in XML format using publicly available Grobid software (v. 0.6.2).

### Phrases

We pre-defined 505 phrases potentially associated with reporting non-significant results (**Suppl. Table 1**). We used a list provided on the Academia Obscura blog, which is based on actual examples found in the biomedical and psychology literature (11).

### Prevalences

We restricted the publication timeframe to three decades: Jan 1990–Sep 2020. The total phrase-positive RCT prevalence was determined for each publication year. To increase the robustness of individual phrase prevalence estimations we binned subsequent RCTs according to their date of the publication into time-periods of three years. For each phrase detected as an exact match in the full texts, time-period prevalences were calculated by dividing the number of RCTs that included one of the 505 phrases describing non-significant results by the total number of RCTs within that period.

### Statistical analysis

To obtain direct evidence on phrase changes over time we used a Bayesian linear regression (12) and determined Bayes factors for each phrase model. This principled ratio measure determines the relative evidence of a model with a linear slope in the temporal prevalence data over a null model with an intercept only. For example, a Bayes factor of 5.0 means that the prevalence of a specific significance phrase over time is five times more probable with a linear change over time than with no linear change over time. Nonetheless, with the tendency of humans to understand the world by applying thresholds to continues spectra, multiple suggestions for interpreting Bayes factor divisions are available. A commonly used list divides the evidence into four strength ranges: Bayes factor between 1–3.2 are ‘not worth more than a bare mention’, between 3.2–10 are ‘substantial’, between 10–100 are ‘strong’ and >100 are ‘decisive’ evidence (13). To our knowledge, there is no evidence that reporting Bayes factors is also subject to suspicious phrasing. We used the R package ‘BayesFactor’ for statistical analysis. Model priors were uninformative.

### Associated P values

Phrases may refer to P values in broadly two types: a direct referral, with the corresponding P value, directly followed after the phrase, mostly in parentheses (E.g., “The drug effect was almost significantly lower in group B (P = 0.052)”). The other type often found in Discussion sections, typically contains longer-range referrals to previously mentioned results, displayed in figures and tables. We tried to quantify the first type of referral by manually extracting the P value within the first 100 characters directly following the extracted phrases within 29,000 random sampled phrases.

## Results

We obtained the full text of 567,758 full-texts of the total of 696,842 PubMed-registered RCTs (81.47%). From the 505 pre-defined significance phrases 272 were at least one time present in the full-text corpus. In total 49,134 RCTs within the 567,758 full texts had a full-text match (61,741 phrases). The yearly prevalences are shown in **Figure 1**. The overall phrase-positive RCT prevalence was stable over time (8.65%, proportional 95% confidence interval: 8.58–8.73%).

**Figure 1.**
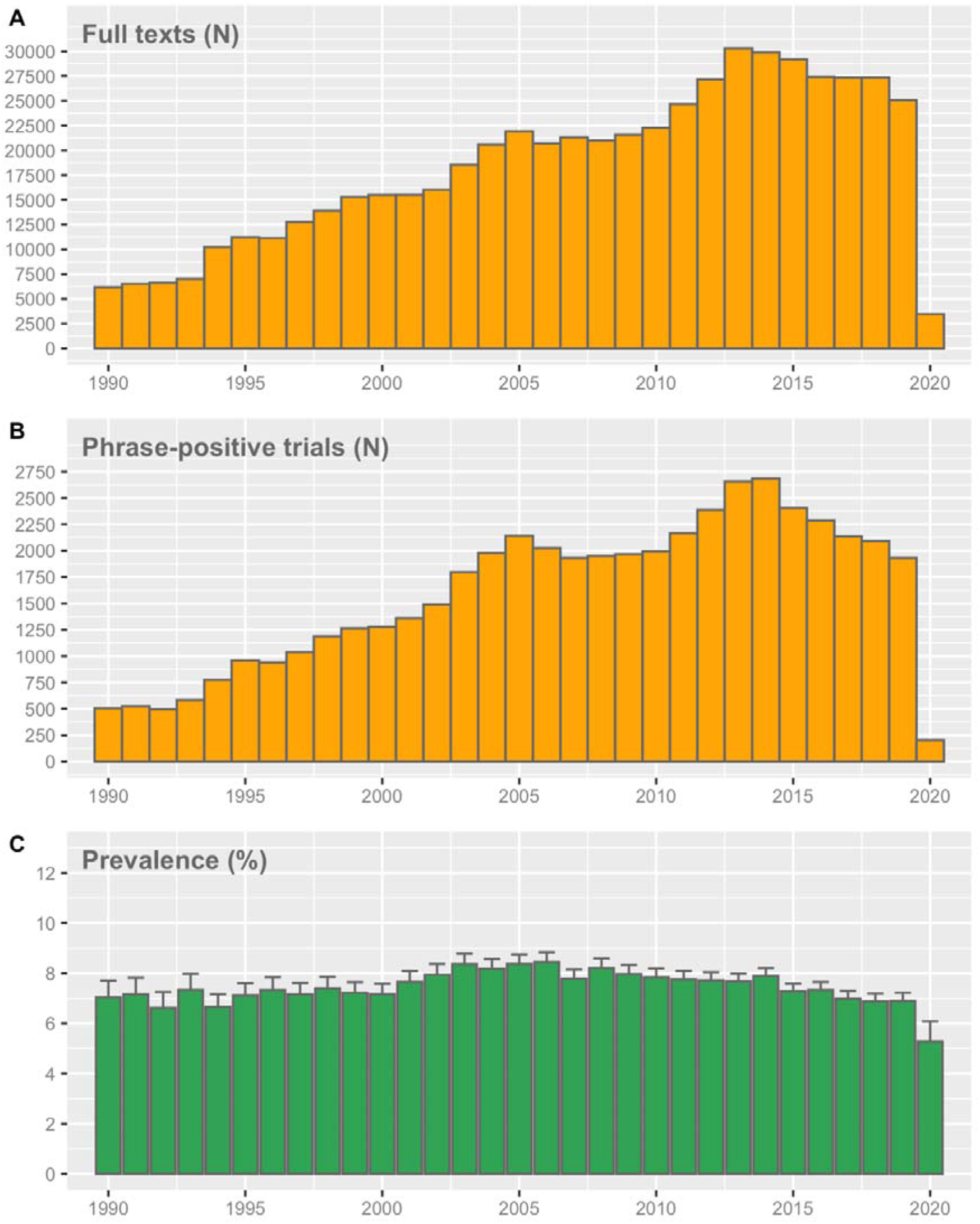
The number of analysed full texts (A), number of phrase-positive RCTs (B) and the corresponding prevalence (C) over time. Error bars represent the proportional 95% confidence interval.

The number of detected RCTs with phrases associated with reporting of non-significant results were unequally distributed (**Table 1**). The most prevalent phrases were ‘marginally significant’ (present in 7,735 RCTs), ‘all but significant’ (7,015 RCTs), ‘a nonsignificant trend’ (3,442 RCTs), ‘failed to reach statistical significance’ (2,578 RCTs) and ‘a strong trend’ (1,700 RCTs).

**Table 1.** The identified number of phrases.

| Phrase | Total RCTs |
| --- | --- |
| marginally significant | 7,735 |
| all but significant | 7,015 |
| a nonsignificant trend | 3,442 |
| failed to reach statistical significance | 2,578 |
| a strong trend | 1,700 |
| nearly significant | 1,391 |
| a clear trend | 1,372 |
| an increasing trend | 1,202 |
| only marginally significant | 1,149 |
| a significant trend | 1,124 |
| potentially significant | 1,104 |
| significant tendency | 1,064 |
| a positive trend | 1,055 |
| a decreasing trend | 962 |
| marginal significance | 887 |
| a slight trend | 885 |
| almost significant | 813 |
| a statistical trend | 811 |
| approaching significance | 796 |
| nominally significant | 740 |
| quite significant | 547 |
| near-significant | 546 |
| an overall trend | 445 |
| likely to be significant | 425 |
| difference was apparent | 409 |
| uncertain significance | 383 |
| did not quite reach statistical significance | 379 |
| a weak trend | 343 |
| marginally statistically significant | 314 |
| tended to be significant | 293 |
| possible significance | 286 |
| not quite significant | 266 |
| a favorable trend | 261 |
| just failed to reach statistical significance | 252 |
| a negative trend | 225 |
| almost reached statistical significance | 219 |
| a possible trend | 218 |
| fell short of significance | 214 |
| not as significant | 204 |
| a small trend | 185 |
| a numerical trend | 184 |
| slightly significant | 182 |
| reached borderline significance | 165 |
| near significance | 156 |
| weakly significant | 147 |
| moderately significant | 146 |
| an apparent trend | 145 |
| barely significant | 135 |
| practically significant | 135 |
| a definite trend | 131 |
| an interesting trend | 129 |
| almost statistically significant | 126 |
| marginally nonsignificant | 101 |
| possibly significant | 100 |
| significantly significant | 100 |
| a marginal trend | 99 |
| close to being significant | 87 |
| just short of significance | 87 |
| fell just short of statistical significance | 77 |
| an obvious trend | 71 |
| tendency toward significance | 66 |
| trending towards significance | 66 |
| a marked trend | 65 |
| a notable trend | 64 |
| at the limit of significance | 62 |
| probably not significant | 62 |
| probably significant | 62 |
| approaches statistical significance | 61 |
| tended toward significance | 59 |
| approached significant | 58 |
| an important trend | 54 |
| marginally insignificant | 53 |
| modestly significant | 53 |
| nearing significance | 49 |
| fairly significant | 47 |
| an observed trend | 46 |
| fell just short of significance | 45 |
| an encouraging trend | 44 |
| not yet significant | 44 |
| not very significant | 42 |
| a borderline significant trend | 40 |
| an unexpected trend | 40 |
| a suggestive trend | 37 |
| a mild trend | 35 |
| a near-significant trend | 34 |
| on the borderline of significance | 31 |
| weak significance | 31 |
| partially significant | 25 |
| strong trend toward significance | 25 |
| approached conventional levels of significance | 24 |
| narrowly missed significance | 24 |
| a strong trend toward significance | 23 |
| not conventionally significant | 22 |
| a pronounced trend | 20 |
| just failed to be significant | 20 |
| tendency toward statistical significance | 20 |
| marginally significant tendency | 19 |
| not highly significant | 19 |
| a slightly increasing trend | 18 |
| an adverse trend | 18 |
| an expected trend | 18 |
| tend to significant | 18 |
| an unfavorable trend | 17 |
| only slightly significant | 17 |
| close to the level of significance | 16 |
| just failed significance | 16 |
| partly significant | 16 |
| barely missed statistical significance | 15 |
| very close to significant | 15 |
| an evident trend | 14 |
| not formally significant | 14 |
| reached near significance | 14 |
| a reliable trend | 13 |
| reasonably significant | 13 |
| on the border of significance | 12 |
| suggestively significant | 12 |
| at the margin of statistical significance | 11 |
| did not quite achieve significance | 11 |
| near to statistical significance | 11 |
| almost insignificant | 10 |
| close to the limit of significance | 10 |
| not fully significant | 10 |
| somewhat significant | 10 |
| a marginal trend toward significance | 9 |
| a statistical trend toward significance | 9 |
| a trend that approached significance | 9 |
| an unexplained trend | 9 |
| just failing to reach statistical significance | 9 |
| not strongly significant | 9 |
| not that significant | 9 |
| slight significance | 9 |
| almost achieved significance | 8 |
| approaching clinical significance | 8 |
| quasi-significant | 8 |
| a worrying trend | 7 |
| almost clinically significant | 7 |
| appeared to be marginally significant | 7 |
| borderline level of statistical significance | 7 |
| not clearly significant | 7 |
| not quite reach the level of significance | 7 |
| not strictly significant | 7 |
| on the margin of significance | 7 |
| on the threshold of significance | 7 |
| trend significance level | 7 |
| very nearly significant | 7 |
| almost attained significance | 6 |
| equivocal significance | 6 |
| possibly statistically significant | 6 |
| probably not statistically significant | 6 |
| scarcely significant | 6 |
| suggestive of statistical significance | 6 |
| weakly statistically significant | 6 |
| a weak trend toward significance | 5 |
| at the edge of significance | 5 |
| just marginally significant | 5 |
| nearly significant tendency | 5 |
| questionably significant | 5 |
| slightly insignificant | 5 |
| a trend close to significance | 4 |
| almost approached significance | 4 |
| bordered on significant | 4 |
| borderline significant trends | 4 |
| indeterminate significance | 4 |
| just borderline significant | 4 |
| just missing significance | 4 |
| on the cusp of significance | 4 |
| on the edge of significance | 4 |
| partial significance | 4 |
| trending towards significant | 4 |
| a distinct trend toward significance | 3 |
| a slight trend toward significance | 3 |
| an associative trend | 3 |
| an elevated trend | 3 |
| an established trend | 3 |
| approached our criterion of significance | 3 |
| approximately significant | 3 |
| arguably significant | 3 |
| better trends of improvement | 3 |
| bordered on being significant | 3 |
| closely significant | 3 |
| did not quite reach conventional levels of statistical significance | 3 |
| effectively significant | 3 |
| fell slightly short of significance | 3 |
| felt short of significance | 3 |
| just missed being statistically significant | 3 |
| not significant by conventional standards | 3 |
| on the verge of significance | 3 |
| verging on significance | 3 |
| virtually significant | 3 |
| a nonsignificant trend toward significance | 2 |
| a numerical increasing trend | 2 |
| a robust trend toward significance | 2 |
| almost significant tendency | 2 |
| approaching but not reaching significance | 2 |
| approaching conventional statistical significance | 2 |
| at the margin of significance | 2 |
| did not quite reach a statistically significant level | 2 |
| fairly close to significance | 2 |
| fell narrowly short of significance | 2 |
| indicative significance | 2 |
| just beyond significance | 2 |
| just escaped significance | 2 |
| just shy of significance | 2 |
| leaning towards significance | 2 |
| narrowly failed significance | 2 |
| nearly reached a significant level | 2 |
| trend bordering on statistical significance | 2 |
| trend in the direction of significance | 2 |
| weakened significance | 2 |
| a certain trend toward significance | 1 |
| a favourable statistical trend | 1 |
| a little significant | 1 |
| a possible trend toward significance | 1 |
| a substantial trend toward significance | 1 |
| a trend significance level | 1 |
| a very slight trend toward significance | 1 |
| almost became significant | 1 |
| almost but not quite significant | 1 |
| approached but fell short of significance | 1 |
| approaching borderline significance | 1 |
| approaching conventional significance levels | 1 |
| approaching marginal significance | 1 |
| approaching, but not reaching, significance | 1 |
| approximating significance | 1 |
| at the limits of significance | 1 |
| at the verge of significance | 1 |
| barely escaped statistical significance | 1 |
| barely insignificant | 1 |
| barely not statistically significant | 1 |
| bordered on being statistically significant | 1 |
| borderline conventional significance | 1 |
| close to the margin of statistical significance | 1 |
| closely not significant | 1 |
| essentially significant | 1 |
| failed to reach significance on this occasion | 1 |
| fell barely short of significance | 1 |
| fell only marginally short of significance | 1 |
| in the verge of significance | 1 |
| just barely failed to reach significance | 1 |
| just fails to reach conventional levels of statistical significance | 1 |
| just outside the bounds of significance | 1 |
| just outside the level of significance | 1 |
| just over the limits of statistical significance | 1 |
| narrowly escaped significance | 1 |
| narrowly missed the significance level | 1 |
| near nominal significance | 1 |
| near-marginal significance | 1 |
| nearly reaching the level of significance | 1 |
| not completely significant | 1 |
| not entirely significant | 1 |
| not especially significant | 1 |
| not globally significant | 1 |
| not markedly significant | 1 |
| not non-significant | 1 |
| not overly significant | 1 |
| not unequivocally significant | 1 |
| only just missed significance at the 5% level | 1 |
| only slightly missed the level of significance | 1 |
| only slightly missed the significance level | 1 |
| rather marginal significance | 1 |
| significant to some degree | 1 |
| slight evidence of significance | 1 |
| slightly missed being of statistical significance | 1 |
| slightly missed statistical significance | 1 |
| slightly not significant | 1 |
| slightly outside the range of significance | 1 |
| technically not significant | 1 |
| tended to approach significance | 1 |
| verged on being significant | 1 |
| very closely brushed the limit of statistical significance | 1 |
| very narrowly missed significance | 1 |
| very slightly significant | 1 |

We found evidence for a temporal change in multiple prevalences (**Suppl. Table 2**). From the phrases with a Bayes factor above 100 the RCT prevalence increased from 0.005 to 0.05% (‘a numerical trend’), 0.098 to 0.23% (‘a positive trend’), 0.067 to 0.346% (‘an increasing trend’) and 0.036 to 0.201% (‘nominally significant’). Whereas the phrases – ‘all but significant’, ‘approaches statistical significance’, ‘did not quite reach statistical significance’, ‘difference was apparent’, ‘failed to reach statistical significance’ and ‘not quite significant’ – sharply decreased over time (**Figure 2**). An additional seventeen phrases had ‘strong’ Bayes factors between 10 and 100 (**Suppl. Figure 1**). Fifteen phrases had a Bayes factor between 3.2 and 10 (**Suppl. Table 2**), indicating ‘substantial’ evidence for a temporal change. The remaining phrases are ‘not worth more than a bare mention’.

**Figure 2.**
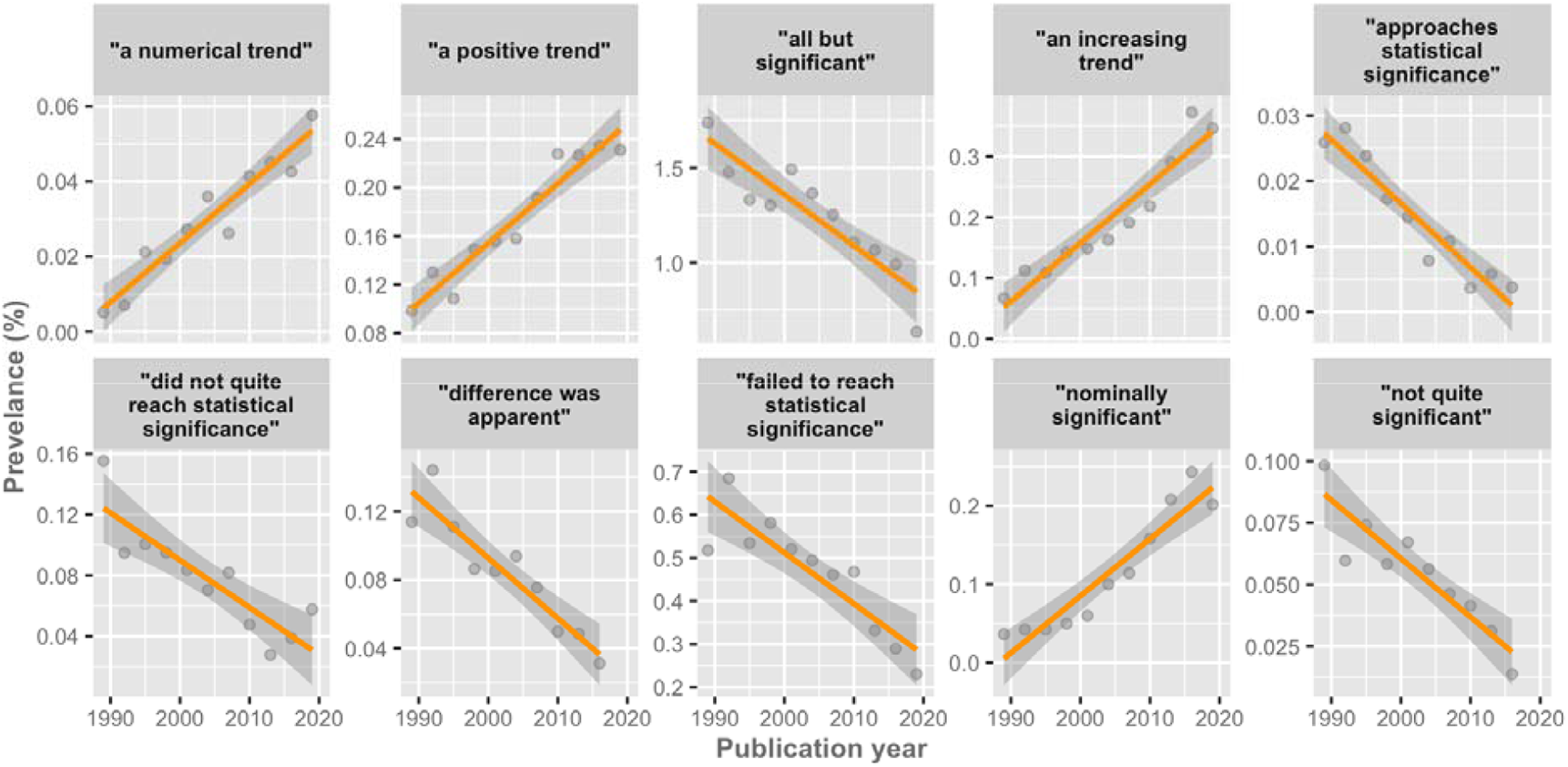
Temporal plots for phrases with ‘decisive’ evidence (i.e., Bayes factors > 100) for temporal change. Prevalence estimates are shown as dots, together with the linear regression model fit and corresponding uncertainty.

### Associated P values

Within the random sample of 29,000 RCTs that contained one of the non-significant phrases, we extracted 11,926 P values (41.1%) that were within the ‘100 characters’ range. Interrater P value variability, based on a sample of fifty similar extractions – hidden within the larger random sample and seen by two authors – was less than 4%.

The P value distribution was characterized with a high prevalence within the 0.05–0.15 range (median: 0.06; 25–75% interval: 0.05–0.08; 5–95% interval: 0.006–0.15; **Figure 3**). The proportions of P values as being categorized as <0.05, between 0.05–0.15 or above 0.15 are given in **Suppl. Table 3**.

**Figure 3.**
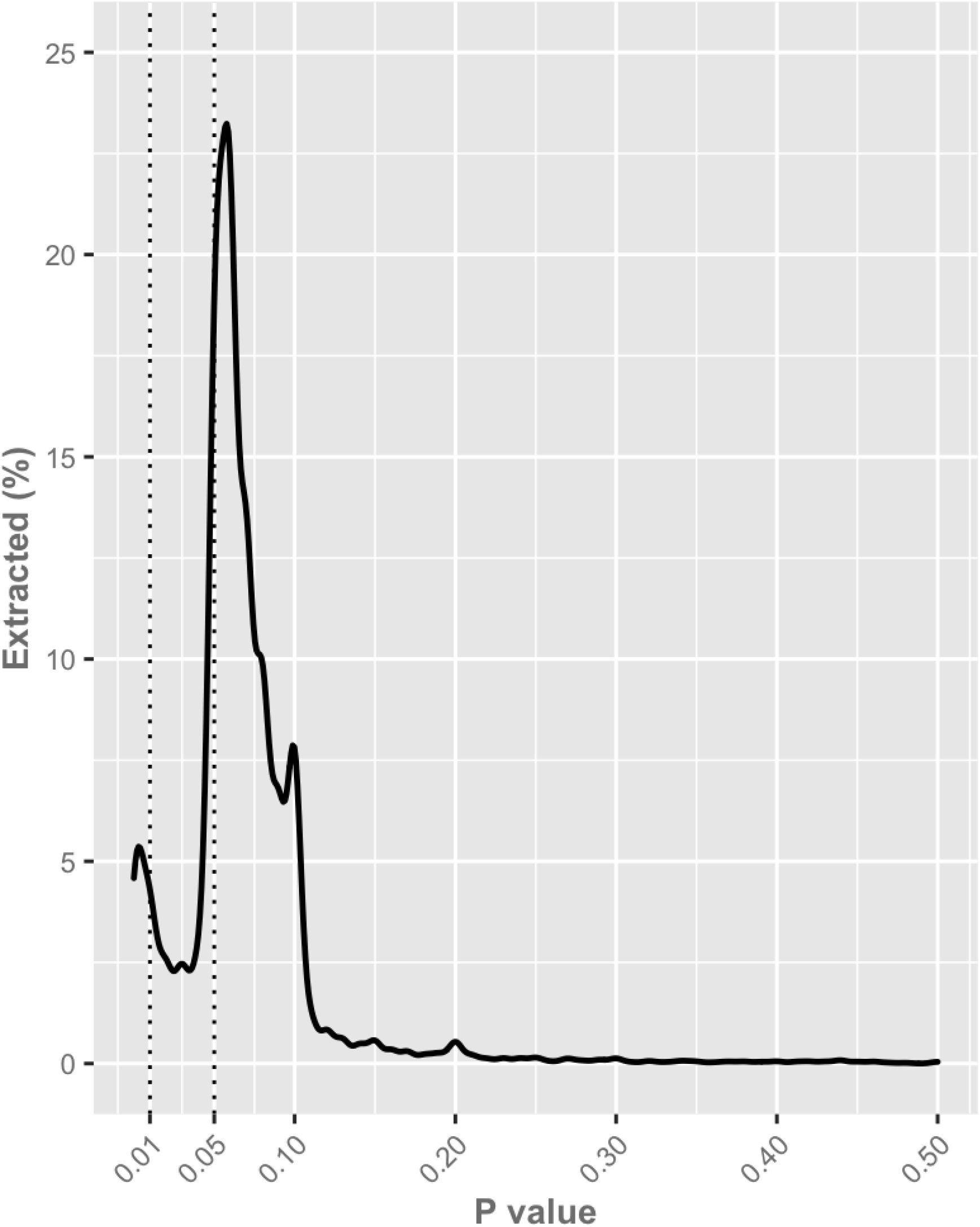
Density plot of the 11,926 manually extracted P values.

Some phrases were highly associated with a P value between 0.05 and 0.15 (**Figure 4** and **Suppl. Figure 2**). Highest percentages for relative frequent phrases were found in this particular range for: “almost reached statistical significance”, “almost significant”, “a strong trend”, “did not quite reach statistical significance”, “just failed to reach statistical significance”, “near significance” and “not quite significant” (**Figure 4**).

**Figure 4.**
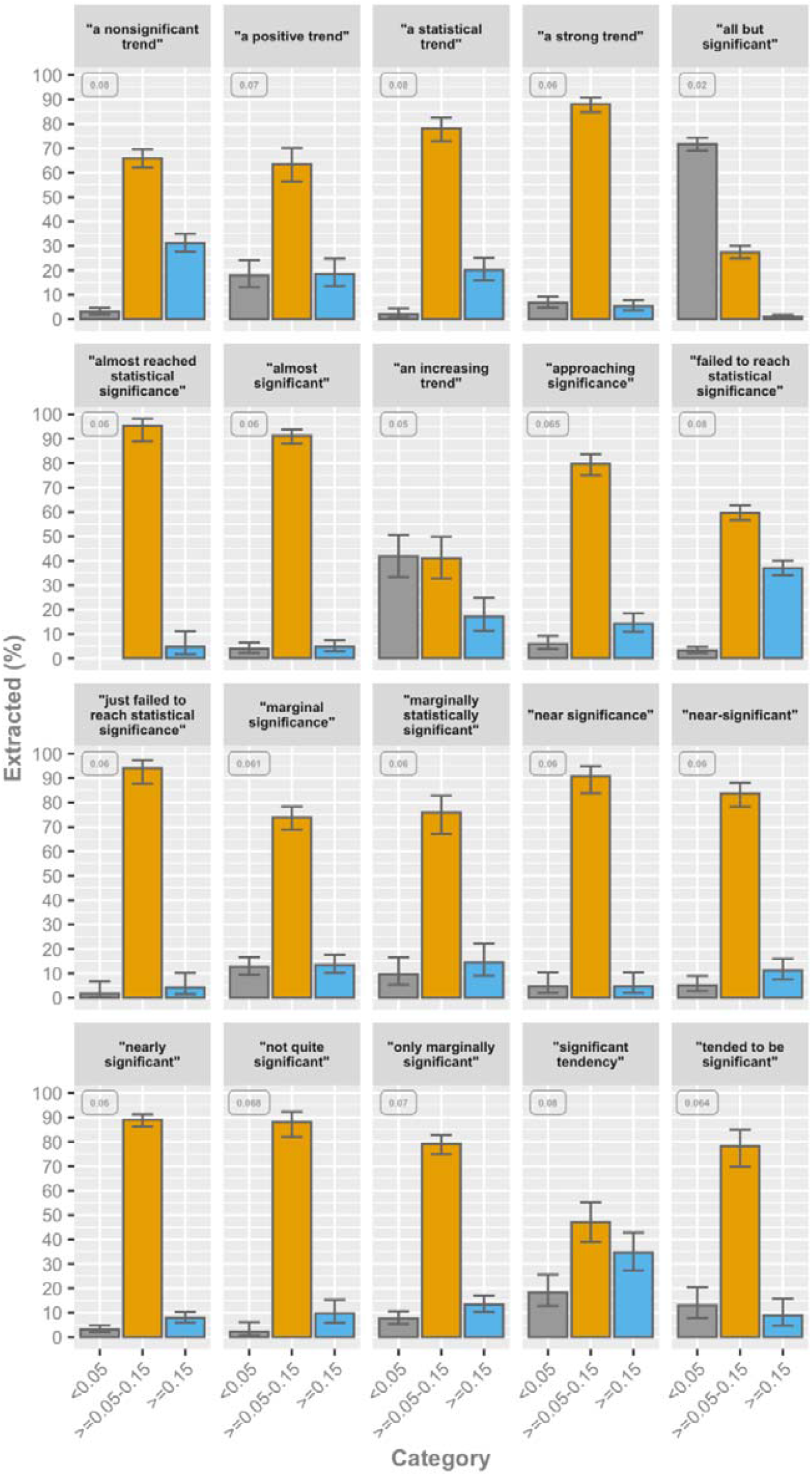
Category percentages for the twenty most frequent phrases describing non-significant results, with at least 100 manually extracted P values. Error bars represent the proportional 95% confidence interval. The associated median P value is presented in the upper left corner of each phrase.

Other phrases were much less linked to 0.05–0.15 P values, namely “a significant trend”, “all but significant”, “an increasing trend” and “nominally significant” (**Figure 4**). For less frequent phrases, similar differences were found between phrases, with some strongly connected to P values just above 0.05 (**Suppl. Figure 2**).

## Discussion

### Principal findings

In this study, we systematically assessed more than half a million RCT full texts on temporal trends and associated P values in phrases linked to non-significant reporting. We present a robust estimate of nine percent of RCTs using specific language to report P values around 0.06. This prevalence remains relatively stable in the past three decades. We also determined fluctuations over time in the frequently used non-significant phrases. Some phrases are gaining popularity over time, whereas others are in decline. In our manual analysis, the vast majority of the phrases describing non-significant results were closely associated with P values in the range of 0.05–0.15.

### Strengths and limitations

This is the first study to explore more than eighty percent of all available PubMed-indexed RCTs on the occurrence of phrases reporting non-significant results. The large sample is required to effectively quantify prevalence and changes in phrasing over time, given the relatively low frequency of several phrases. Moreover, we also quantified the actual P values of the most frequently used phrases reporting non-significant results.

Our study also has inherent limitations. First, we pre-defined more than five hundred phrases denoting results that do not reach formal statistical significance. We may have missed phrases with similar meaning. This would lead to an underestimated overall prevalence. Second, not all phrases are equally specific in their association with P values just above 0.05. Third, we studied English-language RCTs only. Generalizations to other languages can therefore not be made. Fourth, we only had access to published full texts. This prevents us from drawing causal conclusions as non-published manuscripts with specific non-significant phrases, which did not undergo a peer-review process, are not available. Connected to that, despite our data collection in September 2020, we missed a relatively large proportion of RCTs published in 2020, rendering our results less stable for the last year. Fifth, we only characterized P values in the direct vicinity of the phrases. Long-range referrals in the text or tables were not included. The association frequencies may hence be conservatively low.

### Interpretation

Our findings suggest that specific phrasing to report non-significant findings are fairly common in RCTs. RCTs are time-and energy-consuming endeavours, and an ‘almost significant’ result, can, therefore, be a disappointing experience that can be softened with phrases that convey some sort of statistical significance. Our elucidation of the characteristics of the most prevalent phrases can help readers, peer reviewers and editors to detect potential spin in manuscripts that overstate or incorrectly interpret their non-significant results. Our results also support the notion that some phrases are becoming more popular.

The detected P value distributions are important in light of the recent discussions to lower the default P value threshold to 0.005 to improve the validity and reproducibility of novel scientific findings (14). P values near 0.05 are highly dependent on sample size and generally provide weak evidence for the alternative hypothesis. This threshold can consequently lead to high probabilities of false-positive reporting. However, replacing the common 0.05 threshold with an even lower arbitrary value is not a definitive solution. Clinical research is diverse and redefining the term ‘statistical significance’ to even less likely outputs, will probably have negative consequences. Lakens et al. (15) therefore suggest that we should abandon a universal cutoff value and associated ‘statistical significance’ phrasing, and allow scholars to judge clinical relevance of RCT results on a case-by-case basis. Based on our data we think that such a personalized approach is beneficial for everyone – especially since it is currently unknown if P value cutoffs as low as 0.005 do indeed lead to lower false positive reporting and will lead to more rigorous clinical evidence. A stricter threshold requires large sample sizes in replication studies – which are hardly conducted - and will probably increase the risk of presenting underpowered clinical results. Moreover, since it is estimated that half of the results of clinical trials are never published (16), mainly due to negative findings, lowering the P value threshold will result in more ‘negative’ studies that remain largely unpublished. Besides, if authors discuss and judge their threshold value transparently and show the clinical relevance, there is no need to tie oneself to a universal P value cutoff. Journal editors and (statistical) reviewers can play an important role in propagating ideas from the so-called ‘new statistics’ strategy, which aims to switch away from null hypothesis significance testing to using effect sizes and cumulation of evidence to explore and determine potential clinical relevance (17-19). Some argue that Bayes Factors should replace the quest for statistical significance. In our analysis, some phrases were associated with BFs that represent ‘decisive evidence’ for temporal changes. It is worth to mention that BFs are considered a good alternative for statistical significance. However, the BFs may be subject to other types of biases and linguistic persuasion as well, so we are not sure whether this would be the solution.

Based on our study, we question the current state of RCT reporting where scholars feel a certain pressure to creatively phrase their non-significant findings to pass as significant in the process of publishing RCT results. Apart from abandoning a universally held threshold, an additional solution may be the two-step submission process that has gained popularity in the past years (20, 21). This entails that an author first submits a version including the introduction and methods. Based on the reviews of this submission a journal provisionally accepts the manuscript. When the data are collected, the authors can finalize their paper with the results and interpretation, irrespective of the publication status.

In conclusion, we recommend RCT researchers, reviewers and journal editors to abandon the focus on formal statistical significance cutoffs to allow for full transparency in borderline significant results. Fifteen years of advocacy to shift away from null hypothesis testing has not yet fully materialized in RCT publications. We hope our study will stimulate researchers to put their creativity to good use in scientific research and abandon the narrow focus on fixed statistical thresholds with its associated phrases.

## Data Availability

Data sharing: All used PubMed IDs, detected phrases, co-text extractions, manually identified P values, and processing scripts are openly shared at:
https://github.com/wmotte/almost_significant (v1.0; http://doi.org/10.5281/zenodo.4313162).

## Footnotes

### Contributors

All the authors contributed substantially to the study conception and design; and the acquisition, analysis, and interpretation of data. All the authors drafted the work and revised it critically for important intellectual content; gave final approval of the version to be published; and agreed to be accountable for all aspects of the work in ensuring that questions related to the accuracy or integrity of any part of the work are appropriately investigated and resolved. WMO is the guarantor.

### Ethics approval

None required.

### Data sharing

All used PubMed IDs, detected phrases, co-text extractions, manually identified P values, and processing scripts are openly shared at: https://github.com/wmotte/almost_significant (v1.0; http://doi.org/10.5281/zenodo.4313162).

