## Supplementary material for "Almost significant: trends and P values in the use of phrases describing marginally significant results in 567,758 randomized controlled trials published between 1990 and 2020"

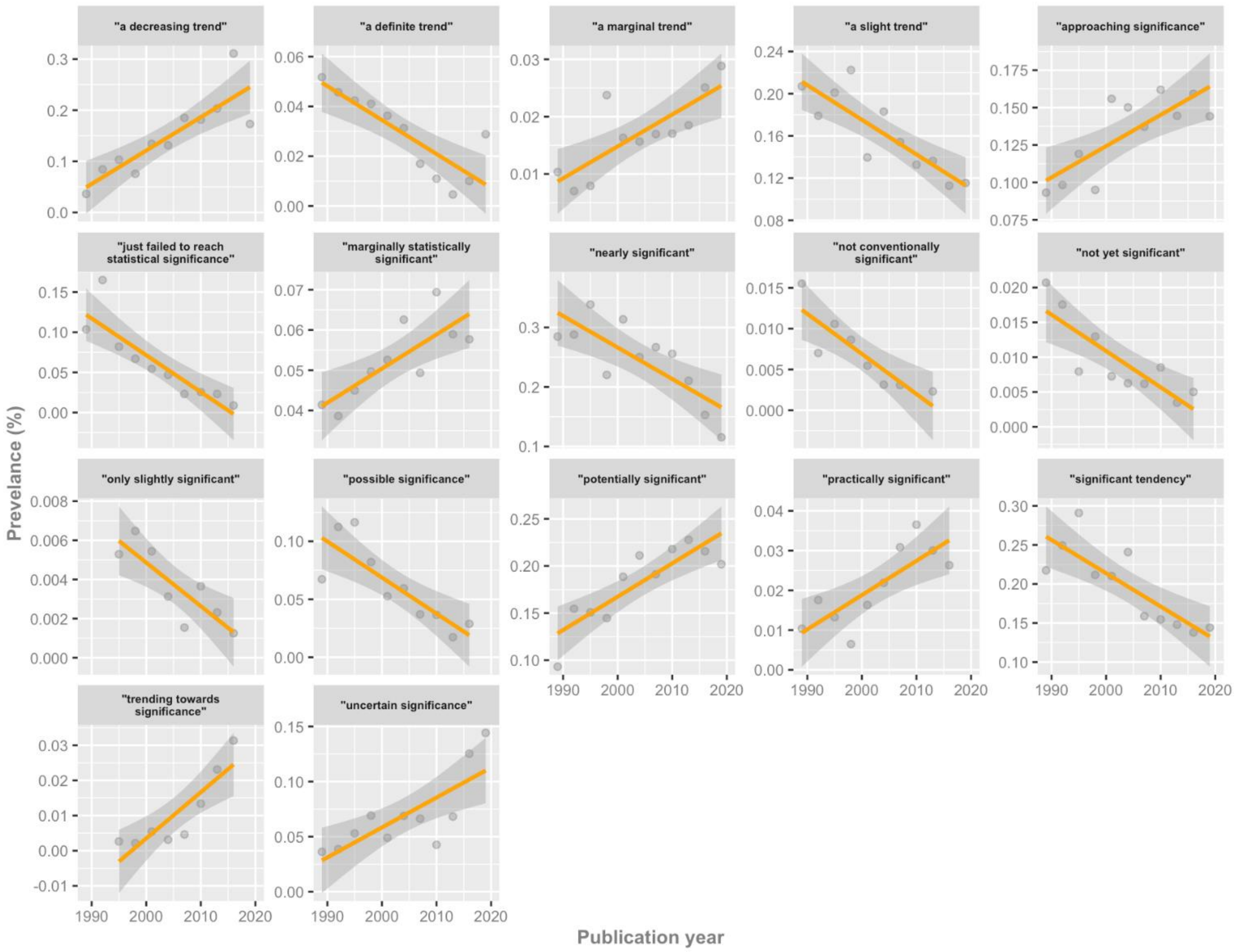


**Figure S1**. Temporal plots for phrases with 'strong' evidence (i.e., Bayes factors between 10–100) for temporal change. Prevalence estimates are shown as dots, together with the linear regression model fit and 95% confidence interval.

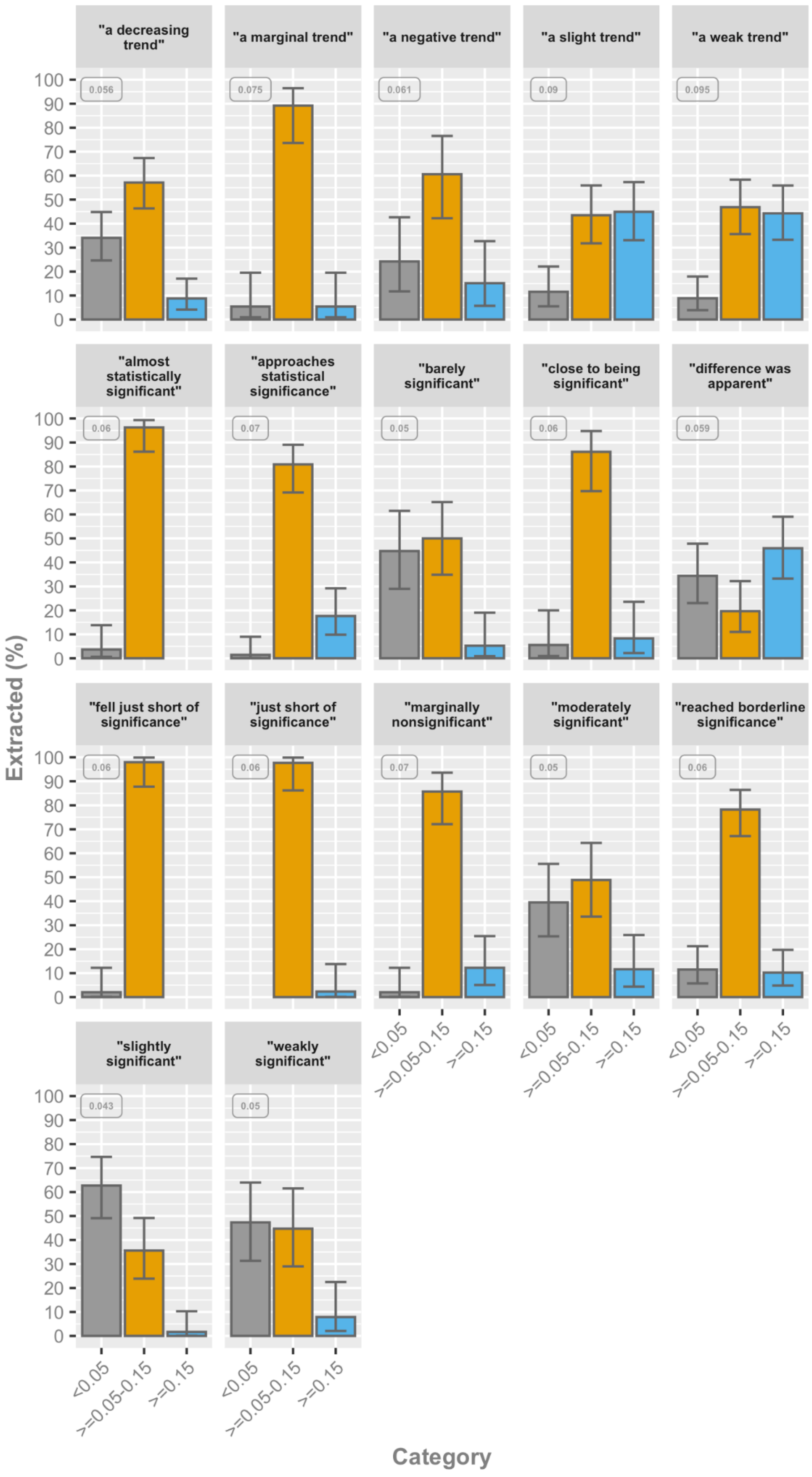


**Figure S2**. Category percentages for the phrases describing non-significant results with the number of manually extracted P values with occurrences between 30 and 100 times in our manual analysis. Error bars represent the proportional 95% confidence interval. The associated median P value is presented in the upper left corner of each phrase

**Table S1**. The 505 pre-defined phrases associated with reporting non-significant results.

| | 1 | 2 | 3 | 4 | 5 |
|---|---|---|---|---|---|
| 1 | barely not statistically significant | approaching an acceptable significance level | fell narrowly short of significance | nearly significant tendency | practically significant |
| 2 | a barely detectable statistically significant difference | approaching borderline significance | fell only marginally short of significance | nearly, but not quite significant | probably not experimentally significant |
| 3 | a borderline significant trend | approaching borderline statistical significance | fell only short of significance | near-marginal significance | probably not significant |
| 4 | a certain trend toward significance | approaching but not reaching significance | fell short of significance | near-significant | probably not statistically significant |
| 5 | a clear tendency to significance | approaching clinical significance | fell slightly short of significance | near-to-significance | probably significant |
| 6 | a clear trend | approaching close to significance | fell somewhat short of significance | near-trend significance | provisionally significant |
| 7 | a clear, strong trend | approaching conventional significance levels | felt short of significance | nominally significant | quasi-significant |
| 8 | a considerable trend toward significance | approaching conventional statistical significance | flirting with conventional levels of significance | non-insignificant result | questionably significant |
| 9 | a decreasing trend | approaching formal significance | heading towards significance | non-significant in the statistical sense | quite close to significance at the 10% level |
| 10 | a definite trend | approaching independent prognostic significance | hint of significance | not absolutely significant but very probably so | quite significant |
| 11 | a distinct trend toward significance | approaching marginal levels of significance | hovered around significance | not as significant | rather marginal significance |
| 12 | a favorable trend | approaching marginal significance | hovered at nearly a significant level | not clearly significant | reached borderline significance |
| 13 | a favourable statistical trend | approaching more closely significance | hovering closer to statistical significance | not completely significant | reached near significance |
| 14 | a little significant | approaching our preset significance level | hovers on the brink of significance | not completely statistically significant | reasonably significant |
| 15 | a margin at the edge of significance | approaching prognostic significance | in the edge of significance | not conventionally significant | remarkably close to significance |
| 16 | a marginal trend | approaching significance | in the verge of significance | not currently significant | resides on the edge of significance |
| 17 | a marginal trend toward significance | approaching the traditional significance level | inconclusively significant | not decisively significant | roughly significant |
| 18 | a marked trend | approaching to statistical significance | indeterminate significance | not entirely significant | scarcely significant |
| 19 | a mild trend | approaching, although not reaching, significance | indicative significance | not especially significant | significant at the .07 level |
| 20 | a moderate trend toward significance | approaching, but not reaching, significance | is just outside the conventional levels of significance | not exactly significant | significant tendency |
| 21 | a near-significant trend | approximately significant | just about significant | not extremely significant | significant to some degree |
| 22 | a negative trend | approximating significance | just above the arbitrary level of significance | not formally significant | significant, or close to significant effects |
| 23 | a nonsignificant trend | arguably significant | just above the margin of significance | not fully significant | significantly significant |
| 24 | a nonsignificant trend toward significance | as good as significant | just at the conventional level of significance | not globally significant | similar but not nonsignificant trends |
| 25 | a notable trend | at the brink of significance | just barely below the level of significance | not highly significant | slight evidence of significance |
| 26 | a numerical increasing trend | at the cusp of significance | just barely failed to reach significance | not markedly significant | slight non-significance |
| 27 | a numerical trend | at the edge of significance | just barely insignificant | not moderately significant | slight significance |
| 28 | a positive trend | at the limit of significance | just barely statistically significant | not non-significant | slight tendency toward significance |
| 29 | a possible trend | at the limits of significance | just beyond significance | not numerically significant | slightly above the level of significance |
| 30 | a possible trend toward | at the margin of | just borderline | not obviously significant | slightly below the level of |

| | | | | | |
|---|---|---|---|---|---|
| | significance | significance | significant | | significance |
| 31 | a pronounced trend | at the margin of statistical significance | just escaped significance | not overly significant | slightly exceeded significance level |
| 32 | a reliable trend | at the verge of significance | just failed significance | not quite borderline significance | slightly failed to reach statistical significance |
| 33 | a robust trend toward significance | at the very edge of significance | just failed to be significant | not quite reach the level of significance | slightly insignificant |
| 34 | a significant trend | barely below the level of significance | just failed to reach statistical significance | not quite significant | slightly less than needed for significance |
| 35 | a slight slide towards significance | barely escaped statistical significance | just failing to reach statistical significance | not quite within the conventional bounds of statistical significance | slightly marginally significant |
| 36 | a slight tendency toward significance | barely escapes being statistically significant at the 5% risk level | just fails to reach conventional levels of statistical significance | not reliably significant | slightly missed being of statistical significance |
| 37 | a slight trend | barely failed to attain statistical significance | just lacked significance | not remarkably significant | slightly missed statistical significance |
| 38 | a slight trend toward significance | barely fails to attain statistical significance at conventional levels | just marginally significant | not significant by common standards | slightly missed the conventional level of significance |
| 39 | a slightly increasing trend | barely insignificant | just missed being statistically significant | not significant by conventional standards | slightly missed the level of statistical significance |
| 40 | a small trend | barely missed statistical significance | just missing significance | not significant by traditional standards | slightly missed the margin of significance |
| 41 | a statistical trend | barely missed the commonly acceptable significance level | just on the verge of significance | not significant in the formal statistical sense | slightly not significant |
| 42 | a statistical trend toward significance | barely outside the range of significance | just outside accepted levels of significance | not significant in the narrow sense of the word | slightly outside conventional statistical significance |
| 43 | a strong tendency towards statistical significance | barely significant | just outside levels of significance | not significant in the normally accepted statistical sense | slightly outside the margins of significance |
| 44 | a strong trend | below but verging on the statistical significant level | just outside the bounds of significance | not significantly significant but clinically meaningful | slightly outside the range of significance |
| 45 | a strong trend toward significance | better trends of improvement | just outside the conventional levels of significance | not statistically quite significant | slightly outside the significance level |
| 46 | a substantial trend toward significance | bordered on a statistically significant value | just outside the level of significance | not strictly significant | slightly outside the statistical significance level |
| 47 | a suggestive trend | bordered on being significant | just outside the limits of significance | not strictly speaking significant | slightly significant |
| 48 | a trend close to significance | bordered on being statistically significant | just outside the traditional bounds of significance | not technically significant | somewhat marginally significant |
| 49 | a trend significance level | bordered on but was not less than the accepted level of significance | just over the limits of statistical significance | not that significant | somewhat short of significance |
| 50 | a trend that approached significance | bordered on significant | just short of significance | not to an extent that was fully statistically significant | somewhat significant |
| 51 | a very slight trend toward significance | borderline conventional significance | just shy of significance | not too distant from statistical significance at the 10% level | somewhat statistically significant |
| 52 | a weak trend | borderline level of statistical significance | just skirting the boundary of significance | not too far from significant at the 10% level | strong trend toward significance |
| 53 | a weak trend toward significance | borderline significant | just tendentially significant | not totally significant | sufficiently close to significance |
| 54 | a worrying trend | borderline significant trends | just tottering on the brink of significance at the 0.05 level | not unequivocally significant | suggestive but not quite significant |
| 55 | all but significant | close to a marginally significant level | just very slightly missed the significance level | not very definitely significant | suggestive of a significant trend |
| 56 | almost achieved significance | close to being significant | leaning towards significance | not very definitely significant from the statistical point of view | suggestive of statistical significance |
| 57 | almost approached significance | close to being statistically significant | leaning towards statistical significance | not very far from significance | suggestively significant |
| 58 | almost attained | close to borderline | likely to be significant | not very significant | tailed to insignificance |

| | | | | | |
|---|---|---|---|---|---|
| | significance | significance | | | |
| 59 | almost became significant | close to the boundary of significance | loosely significant | not very statistically significant | tantalisingly close to significance |
| 60 | almost but not quite significant | close to the level of significance | marginal significance | not wholly significant | technically not significant |
| 61 | almost clinically significant | close to the limit of significance | marginally and negatively significant | not yet significant | teetering on the brink of significance |
| 62 | almost insignificant | close to the margin of significance | marginally insignificant | not strongly significant | tend to significant |
| 63 | almost marginally significant | close to the margin of statistical significance | marginally nonsignificant | noticeably significant | tended to approach significance |
| 64 | almost non-significant | closely approaches the brink of significance | marginally outside the level of significance | on the border of significance | tended to be significant |
| 65 | almost reached statistical significance | closely approaches the statistical significance | marginally significant | on the borderline of significance | tended toward significance |
| 66 | almost significant | closely approximating significance | marginally significant tendency | on the borderlines of significance | tendency toward significance |
| 67 | almost significant tendency | closely not significant | marginally statistically significant | on the boundaries of significance | tendency toward statistical significance |
| 68 | almost statistically significant | closely significant | may not be significant | on the boundary of significance | tends to approach significance |
| 69 | an adverse trend | close-to-significant | medium level of significance | on the brink of significance | tentatively significant |
| 70 | an apparent trend | did not achieve conventional threshold levels of statistical significance | mildly significant | on the cusp of conventional statistical significance | too far from significance |
| 71 | an associative trend | did not exceed the conventional level of statistical significance | missed narrowly statistical significance | on the cusp of significance | trend bordering on statistical significance |
| 72 | an elevated trend | did not quite achieve acceptable levels of statistical significance | moderately significant | on the edge of significance | trend in a significant direction |
| 73 | an encouraging trend | did not quite achieve significance | modestly significant | on the limit to significant | trend in the direction of significance |
| 74 | an established trend | did not quite achieve the conventional levels of significance | narrowly avoided significance | on the margin of significance | trend significance level |
| 75 | an evident trend | did not quite achieve the threshold for statistical significance | narrowly eluded statistical significance | on the threshold of significance | trending towards significance |
| 76 | an expected trend | did not quite attain conventional levels of significance | narrowly escaped significance | on the verge of significance | trending towards significant |
| 77 | an important trend | did not quite reach a statistically significant level | narrowly evaded statistical significance | on the very borderline of significance | uncertain significance |
| 78 | an increasing trend | did not quite reach conventional levels of statistical significance | narrowly failed significance | on the very fringes of significance | vaguely significant |
| 79 | an interesting trend | did not quite reach statistical significance | narrowly missed achieving significance | on the very limits of significance | verged on being significant |
| 80 | an inverse trend toward significance | did not reach the traditional level of significance | narrowly missed overall significance | only a little short of significance | verging on significance |
| 81 | an observed trend | did not reach the usually accepted level of clinical significance | narrowly missed significance | only just failed to meet statistical significance | verging on the statistically significant |
| 82 | an obvious trend | difference was apparent | narrowly missed standard significance levels | only just insignificant | verging-on-significant |
| 83 | an overall trend | direction heading towards significance | narrowly missed the significance level | only just missed significance at the 5% level | very close to approaching significance |
| 84 | an unexpected trend | does not appear to be sufficiently significant | narrowly missing conventional significance | only marginally fails to be significant at the 95% level | very close to significant |
| 85 | an unexplained trend | does not narrowly reach statistical significance | near limit significance | only marginally nearly insignificant | very close to the conventional level of significance |
| 86 | an unfavorable trend | does not reach the conventional | near miss of statistical significance | only marginally significant | very close to the cut-off for significance |

| | | | | | |
|---|---|---|---|---|---|
| | | significance level | | | |
| 87 | appeared to be marginally significant | effectively significant | near nominal significance | only slightly less than significant | very close to the established statistical significance level of p=0.05 |
| 88 | approached acceptable levels of statistical significance | equivocal significance | near significance | only slightly missed the conventional threshold of significance | very close to the threshold of significance |
| 89 | approached but did not quite achieve significance | essentially significant | near to statistical significance | only slightly missed the level of significance | very closely approaches the conventional significance level |
| 90 | approached but fell short of significance | extremely close to significance | near significance | only slightly missed the significance level | very closely brushed the limit of statistical significance |
| 91 | approached conventional levels of significance | failed to reach significance on this occasion | near-borderline significance | only slightly non-significant | very narrowly missed significance |
| 92 | approached near significance | failed to reach statistical significance | near-certain significance | only slightly significant | very nearly significant |
| 93 | approached our criterion of significance | fairly close to significance | nearing significance | partial significance | very slightly non-significant |
| 94 | approached significant | fairly significant | nearly acceptable level of significance | partially significant | very slightly significant |
| 95 | approached the borderline of significance | falls just short of standard levels of statistical significance | nearly approaches statistical significance | partly significant | virtually significant |
| 96 | approached the level of significance | fell just short of significance | nearly borderline significance | perceivable statistical significance | weak significance |
| 97 | approached trend levels of significance | fell barely short of significance | nearly negatively significant | possible significance | weakened significance |
| 98 | approached, but did reach, significance | fell just short of significance | nearly positively significant | possibly marginally significant | weakly non-significant |
| 99 | approaches but fails to achieve a customary level of statistical significance | fell just short of statistical significance | nearly reached a significant level | possibly significant | weakly significant |
| 100 | approaches statistical significance | fell just short of the traditional definition of statistical significance | nearly reaching the level of significance | possibly statistically significant | weakly statistically significant |
| 101 | approaching a level of significance | fell marginally short of significance | nearly significant | potentially significant | well-nigh significant |

**Table S2**. The evidence of temporal change in the phrases with at least five time-points expressed as the Bayes factor relative to no temporal change (lower threshold set to 2.0). The colours represent the strength of evidence as specified in the main text.

| Bayes factor | Phrase |
|---|---|
| 9023.2 | "a positive trend" |
| 5970.0 | "a numerical trend" |
| 4381.4 | "an increasing trend" |
| 1990.3 | "nominally significant" |
| 1316.2 | "approaches statistical significance" |
| 363.9 | "difference was apparent" |
| 338.0 | "all but significant" |
| 198.3 | "not quite significant" |
| 158.5 | "failed to reach statistical significance" |
| 110.4 | "did not quite reach statistical significance" |
| 77.9 | "a decreasing trend" |
| 71.8 | "potentially significant" |
| 67.2 | "a slight trend" |
| 64.0 | "just failed to reach statistical significance" |

| | |
|---|---|
| 50.9 | "a definite trend" |
| 39.5 | "significant tendency" |
| 27.6 | "not yet significant" |
| 25.4 | "possible significance" |
| 24.0 | "a marginal trend" |
| 21.1 | "nearly significant" |
| 19.7 | "approaching significance" |
| 17.8 | "trending towards significance" |
| 17.4 | "uncertain significance" |
| 16.0 | "not conventionally significant" |
| 15.5 | "practically significant" |
| 15.0 | "marginally statistically significant" |
| 11.3 | "only slightly significant" |
| 10.0 | "a statistical trend" |
| 7.6 | "probably not significant" |
| 7.0 | "a negative trend" |
| 6.8 | "barely significant" |
| 5.8 | "an overall trend" |
| 5.4 | "slightly significant" |
| 4.9 | "a reliable trend" |
| 4.5 | "almost significant" |
| 4.2 | "tended to be significant" |
| 4.0 | "almost achieved significance" |
| 3.9 | "trend significance level" |
| 3.8 | "on the borderline of significance" |
| 3.7 | "almost insignificant" |
| 3.5 | "tendency toward significance" |
| 3.2 | "a favorable trend" |
| 3.1 | "not quite reach the level of significance" |
| 2.9 | "almost statistically significant" |
| 2.9 | "moderately significant" |
| 2.9 | "approached conventional levels of significance" |
| 2.8 | "not highly significant" |
| 2.8 | "slight significance" |
| 2.7 | "a notable trend" |
| 2.7 | "just failing to reach statistical significance" |
| 2.7 | "marginally nonsignificant" |
| 2.5 | "just failed significance" |
| 2.5 | "an observed trend" |
| 2.5 | "possibly significant" |
| 2.4 | "scarcely significant" |

| | |
|---|---|
| 2.3 | "at the margin of statistical significance" |
| 2.3 | "just failed to be significant" |
| 2.3 | "an unexplained trend" |
| 2.3 | "barely missed statistical significance" |
| 2.2 | "weak significance" |
| 2.2 | "modestly significant" |
| 2.0 | "fairly significant" |
| 2.0 | "not fully significant" |
| 2.0 | "likely to be significant" |
| 2.0 | "almost attained significance" |
| 2.0 | "a statistical trend toward significance" |

**Table S3**. All extracted P values within the three range categories, as the proportion of the total of 11,926 extractions.

| Category | K | proportion | 95% confidence interval |
|---|---|---|---|
| <0.05 | 2052 | 0.172 | 0.165-0.179 |
| >=0.05-0.15 | 8126 | 0.681 | 0.673-0.69 |
| >=0.15 | 1748 | 0.147 | 0.14-0.153 |
